# Is chronic pain an affective disorder? Moderation of resting-state functional connectivity within the emotional brain by depressive symptoms

**DOI:** 10.1101/2022.02.11.22270877

**Authors:** Yann Quidé, Nell Norman-Nott, Negin Hesam-Shariati, James H. McAuley, Sylvia M. Gustin

**Affiliations:** NeuroRecovery Research Hub, School of Psychology, The University of New South Wales (UNSW), Sydney, NSW, Australia; Centre for Pain IMPACT, Neuroscience Research Australia, Randwick, NSW, Australia; School of Health Sciences, The University of New South Wales (UNSW), Sydney, NSW, Australia

**Keywords:** chronic pain, depression, functional connectivity, emotion, mood

## Abstract

**Background:** Depressive symptoms are often comorbid to chronic pain. These conditions share aberrant emotion processing and regulation, as well as common brain networks. However, the relationship between depressive symptoms and chronic pain on emotional brain function is unclear.

**Methods:** Participants were 26 individuals with chronic pain (referred to as the Pain group) and 32 healthy controls (HC), who underwent resting-state functional magnetic resonance imaging and completed the Beck Depressive Inventory. Main effects of group, depressive symptom severity (total score), and their interaction were evaluated on functional connectivity from three seed regions (separately, the left and right amygdalae, the medial prefrontal cortex, mPFC) and the rest of the brain. In case of significant interaction, moderation analyses were conducted.

**Results:** The group-by-depressive symptoms interaction was significantly associated with changes in connectivity between the right amygdala and the mPFC (*pFWEc*<0.001). Moderation analysis indicated that, compared to the HC group, the Pain group showed weaker connectivity between these regions at lower levels of depressive symptoms (*p*=0.018), and stronger connectivity at higher levels of depressive symptoms (*p*=0.001). In addition, the strength of connectivity decreased in the HC (*p*=0.004) and increased in the Pain group (*p*=0.011) as the severity of depressive symptoms increased. Finally, the Pain group showed significant weaker connectivity between the mPFC seed and the left parahippocampal gyrus compared to the HC group (*pFWEc*=0.015).

**Conclusions:** These results indicate that depressive symptoms moderate the impact of chronic pain on the emotional brain function and highlight potential implications for the choice of treatment for chronic pain.

## Introduction

Chronic pain is defined as persistent experience of pain for over a period of three months, beyond normal healing time after injury or illness (Treede *et al*., 2015). More specifically, chronic neuropathic pain is a consequence of nerve injury, insult or disease, and is a major health problem, occurring in around 20% of the general population (Breivik *et al*., 2006), with very limited successful therapeutic options. Along with anticonvulsants (gabapentine, pregabaline), antidepressants, including serotonin noradrenaline reuptake inhibitors (SNRIs) or tricyclic antidepressants, are considered first line recommendations by the International Association for the Study of Pain (Finnerup *et al*., 2015). This may be due to, at least in part, aberrant affective and mood processing and regulation being common comorbid consequences of chronic pain (Bair *et al*., 2003). Although depression or depressive symptoms being reported in around 60% of chronic pain sufferers (Bair *et al*., 2003, Rayner *et al*., 2016), the relationship between chronic pain and depression on brain function is not well characterised. In particular, it remains unclear how both conditions impact shared brain phenotypes, and if the impact of depression is additive to the features reported in chronic pain. The present study aimed to determine the independent and interactive effects of chronic pain and depressive symptoms on emotional brain function.

Resting-state functional magnetic resonance imaging is used to identify patterns of functional connectivity within and between brain networks. At rest, the so-called default mode network (DMN), including the medial prefrontal cortex (mPFC), the posterior cingulate cortex (PCC)/precuneus, the left and right temporo-parietal junctions and subcortical regions such as the hippocampus/hippocampal formation (Buckner *et al*., 2008, Greicius *et al*., 2003, Huijbers *et al*., 2011, Raichle *et al*., 2001), is strongly activated and anti-correlated to the ‘de-activated’ central-executive network (CEN), made of the bilateral dorsolateral prefrontal cortex and inferior parietal lobules (Seeley *et al*., 2007). According to the triple network model of psychopathology (Menon, 2011), the salience network, made of the dorsal anterior cingulate cortex, bilateral anterior insula/inferior frontal gyri and subcortical regions such as the amygdala or the nucleus accumbens (Menon, 2015, Uddin, 2015), is responsible for switching brain states from cognitively engaged (activated CEN, deactivated DMN) to rest (CEN deactivated, DMN activated) and vice-versa when necessary.

Transition to chronic pain is associated with changes of connectivity within and between these networks (Pfannmoller and Lotze, 2019). In particular, stronger connectivity between the mPFC and the nucleus accumbens (Baliki *et al*., 2012) is associated with the chronification of pain. When comparing individuals transitioning to chronic pain to recovering individuals, further graph theoretical approaches refined this model, showing increased density of connectivity of a network including the dorsal mPFC, nucleus accumbens and the amygdala (Vachon-Presseau *et al*., 2016). More generally, subcortical (amygdala, hippocampus, nucleus accumbens) and cortical regions (mPFC) involved in emotion processing, affect regulation and memory have been consistently associated with pain experience (Martucci and Mackey, 2018, Ochsner *et al*., 2006, Schmidt *et al*., 2016).

Individuals diagnosed with major depressive disorder also show aberrant connectivity between these regions. Compared to healthy individuals, people with major depressive disorder show decreased functional connectivity within the DMN, especially between the mPFC and PCC/precuneus nodes (Yan *et al*., 2019), as well as between the amygdala and the mPFC (Connolly *et al*., 2017, Tang *et al*., 2013, Veer *et al*., 2010). In addition, further evidence indicates that compared to healthy individuals, people with major depressive disorder show weak positive connectivity between the nucleus accumbens and the mPFC, and strong negative connectivity between the amygdala and the mPFC (Kaiser *et al*., 2015). Although depressive symptoms and chronic pain often co-occur and may share aberrant neurochemical balance and brain networks (Sheng *et al*., 2017), it is also possible that the severity of depressive symptoms may moderate the changes in functional connectivity between regions involved in emotion processing and/or regulation associated with chronic pain.

In this study, we set out to determine whether the severity of depressive symptoms reported moderated the changes in functional connectivity with regions involved in emotion processing (amygdala) and regulation (mPFC), among people suffering from chronic pain and healthy individuals. In particular, consistent with patterns observed in major depressive disorder (Connolly *et al*., 2017, Tang *et al*., 2013, Veer *et al*., 2010), we expected that increasing levels of depressive symptoms would be associated with weaker amygdala-mPFC connectivity in healthy participants, and with stronger connectivity in people with chronic pain (Vachon-Presseau *et al*., 2016). In addition, we expected that depressive symptoms would be associated with aberrant connectivity within the DMN, particularly between the mPFC and the PCC/precuneus, independently of the group (Yan *et al*., 2019).

## Methods

All participants were volunteers who provided informed consent according to procedures approved by the Human Research Ethics committees of the University of New South Wales (HC15206), the University of Sydney (HREC06287) and Northern Sydney Local Health District (1102-066M). The authors assert that all procedures contributing to this work comply with the ethical standards of the relevant national and institutional committees on human experimentation and with the Helsinki Declaration of 1975, as revised in 2008.

### Participants

Participants were 26 individuals suffering from chronic pain (together referred to as the Pain group), including spinal cord injury neuropathic pain (n=11), painful temporomandibular disorder (TMD, n=4), trigeminal neuropathic pain (TNP, n=10), postherpetic neuralgia (n=1), and 32 pain-free healthy controls (HC; see details Table 1). All participants in the Pain group experienced chronic pain, that is pain greater than 3 months (International Association for the Study of Pain, 1986). Neuropathic pain after spinal cord injury was diagnosed according to the International Association for the Study of Pain Spinal Cord Injury Pain Taxonomy (Bryce *et al*., 2012). All people with spinal cord injury suffered from a complete paraplegia with continuous burning and/or shooting pain in areas of sensorimotor loss. Painful TMD is characterised by ongoing musculoskeletal facial pain as assessed using the Research Diagnostic Criteria for TMD (Dworkin and LeResche, 1992). TNP and postherpetic neuralgia, both characterised by continuous dull neuropathic facial pain with sharp exacerbations (Paterno and Singla, 2014), were diagnosed using the Liverpool Criteria (Nurmikko and Eldridge, 2001).

**Table 1.** Sociodemographic and clinical characteristics of the studied cohort

|  | HC<br>(N=32) | Pain<br>(N=26) | Statistics<br>*Welch/t/χ<sup>2</sup>* | *df* | *p* |
| --- | --- | --- | --- | --- | --- |
| Age, mean (SD) [range] | 45.29 (15.78) [22.19-71.76] | 51.64 (10.28) [23.77-79.18] | 1.846 | 53.67 | 0.070 |
| Sex, n (F/M) | 16/16 | 15/11 | 0.341 | 1 | 0.559 |
| BDI total score (SD) [range] | 5.09 (4.63) [0-19] | 13.30 (2.67) [0-34] | **2.325** | **38.30** | **0.025** |
| No or minimal depression (BDI score ≤ 9), N (%) | 29 (91%) | 17 (65%) |  |  |  |
| Mild depression (10 ≤ BDI score ≤ 18), N (%) | 2 (6%) | 6 (23%) |  |  |  |
| Moderate depression (19 ≤ BDI score ≤ 29), N (%) | 1 (3%) | 2 (8%) |  |  |  |
| Severe depression (BDI score ≥ 30), N (%) | 0 | 1 (4%) |  |  |  |
| STAI State, mean (SD) [range] | 27.88 (8.40) [20-54] | 31.42 (9.30) [14-46] | 1.525 | 56 | 0.133 |
| STAI Trait, mean (SD) [range]* | 31.16 (8.19) [20-50] | 36.04 (10.92) [19-53] | 1.925 | 55 | 0.059 |
| Pain duration, mean (SD) [range] | - | 14.04 (12.80) [2-48] | - | - | - |
| VAS pain diary, mean (SD) [range] | - | 3.90 (2.04) [0.2-8.8] | - | - | - |
| VAS scan Pain, mean (SD) [range] | - | 3.03 (1.87) [0-6.6] | - | - | - |
| Medication, n (%) | - | 11 (42%) | - | - | - |
| Amitriptyline, n (%) | - | 1 (4%) | - | - | - |
| Gabapentine, n (%) | - | 2 (8%) | - | - | - |
| Pregabalin, n (%) | - | 3 (12%) | - | - | - |
| Desvenlafaxine (SNRI), n (%) | - | 1 (4%) | - | - | - |
| Paracetamol PRN, n (%) | - | 1 (4%) | - | - | - |
| Pregabalin + oxycodone, n (%) | - | 1 (4%) | - | - | - |
| Oxycodone + paracetamol PRN, n (%) | - | 1 (4%) | - | - | - |
| Codeine + Paracetamol + Ibuprofen PRN, n (%) | - | 1 (4%) | - | - | - |
| Pain location |  |  |  |  |  |
| Left, n (%) | - | 3 (12%) | - | - | - |
| Right, n (%) | - | 4 (15%) | - | - | - |
| Bilateral, n (%) | - | 19 (73%) | - | - | - |

HC: healthy controls; Pain: individuals with chronic pain; df: degrees of freedom; SD: standard deviation; F/M: females/males; BDI: Beck Depression Inventory; STAI: State/Trait Anxiety Inventory; VAS: visual analogue scale; SNRI: selective serotonin and norepinephrine reuptake inhibitor; PRN: Pro re nata
\* Missing value for one HC participant
Significant group differences are in bold

### Assessments

Severity of depressive symptoms were measured using the sum of all 21 items (BDI total score, ranging from 0-63) from the Beck Depression Inventory (BDI) (Beck *et al*., 1961). Cut-off scores for estimation of the severity of depressive symptoms are: 0 to 9 for no or minimal depression, 10 to 18 for mild depression, 19 to 29 for moderate depression, and 30 to 63 for severe depression (Beck *et al*., 1988). The BDI is a reliable measure of depressive symptoms in chronic pain populations (Geisser *et al*., 1997). Severity of state and trait anxiety were assessed using the two 20-item subscales (scores ranging 20-80) from the State-Trait Anxiety Inventory (STAI) (Spielberger *et al*., 1983). Pain intensity was measured in the Pain group only using a visual analogue scale (VAS): participants reported their experienced levels of pain on a 10-cm horizontal ruler, with “no pain” being at the beginning of the ruler (0cm mark), and “worst pain imaginable” at the other extremity (10cm mark) three times a day (morning, noon and evening). Two measures of pain intensity were collected using the VAS: the “pain diary” consists of an average measure of pain intensity for seven days prior to the scanning day, and the “scan pain” is a retrospective measure of pain intensity while the participant was lying in the scanner. Duration of pain, pain location (left, right or both sides of the body), and medications used were also recorded (see Table 1).

### Magnetic resonance imaging

Imaging data were acquired for each participant on two Philips 3T Achieva TX scanner (Philips Healthcare, The Netherlands) housed at Neuroscience Research Australia (Randwick, NSW, Australia; nHC=16, nPain=12) or at St Vincent’s Hospital (Darlinghurst, NSW, Australia; nHC=16, nPain=14). Both scanners were equipped with 8-channel head-coils and used the same acquisition parameters to collect a 3D T1-weighted structural image covering the entire brain: repetition time (TR) = 5.6ms, echo time (TE) = 2.5ms, field of view = 250 × 250 × 174mm, matrix 288 × 288, 200 sagittal slices, flip angle = 8°, voxel size 0.9 × 0.9 × 0.9mm. Furthermore, 180 whole brain T2* weighted echo-planar images (EPI) were acquired (TR=2000ms, TE=30ms, field of view 240 × 140 × 240mm, matrix 80 × 78, 35 slices, slice thickness = 4mm, flip angle = 90°, voxel size 3 × 3 × 4mm) were acquired, with participants asked to close their eyes and let their mind wander without falling asleep.

Pre-processing was performed with the CONN toolbox (v20b; https://sites.google.com/view/conn/) (Whitfield-Gabrieli and Nieto-Castanon, 2012) for SPM12 (v7771, Wellcome Department of Cognitive Neurology, University College London, UK; https://www.fil.ion.ucl.ac.uk/spm/) in Matlab r2021a (Matworks Inc., Sherborn, MA, USA). In addition to automatically discard dummy scans, the first five acquisitions (10s) for each subject were tagged as “outliers” during the CONN outlier detection step and their effect were removed during the denoising step. Pre-processing steps included realignment and unwarping, outlier slices identification (ART toolbox, with movement translation threshold: 0.9 mm and signal intensity threshold: z = 5), segmentation and normalization of the functional and structural images, smoothing with a 8 mm Gaussian kernel. Temporal band-pass filtering (0.008 < f < 0.09) was applied to reduce the effects of low-frequency drift and high-frequency noise. As per the defaults settings of the toolbox, physiological and other potential sources of noise (white matter, cerebro-spinal fluid) were estimated using a component-based noise correction (CompCor) method (Behzadi *et al*., 2007) and regressed out along with movement-related (Friston *et al*., 1996), the constant and first-order linear session effects, and scrubbing (Power *et al*., 2014) covariates. Only participants with less than 18 volumes (10% of the total number of acquisitions) identified as outliers by ART (other than the 5 initial volumes) were considered and included in the analyses.

### Seed-based connectivity

Seed-based functional connectivity maps (seed-to-voxel bivariate correlations) were derived for all participants and all ROIs available within the CONN toolbox. The mPFC seed region was from the CONN network cortical ROIs atlas (Whitfield-Gabrieli and Nieto-Castanon, 2012), derived from an independent component analysis performed on 497 participants from the Human Connectome Project dataset (10□mm diameter spheres around MNI coordinates [1,55,-3]), and the left and right amygdala seed regions were from the FSL Harvard-Oxford maximum likelihood subcortical atlas (HarvardOxford-sub-maxprob-thr25-1mm.nii). Individual functional connectivity maps (bivariate correlations) for these seed regions were then exported for further group-level analyses in SPM12.

### Harmonization

Before entering second-level (group) analyses, individual first-level connectivity maps (for each seed region) were harmonized using the python-based *neuroHarmonize* tools (https://github.com/rpomponio/neuroHarmonize) (Pomponio *et al*., 2020). Briefly, this approach uses empirical Bayes methods derived from the ‘ComBat’ R package (Johnson *et al*., 2007) to adjust whole-brain statistical maps for variation associated with scanning location in multi-site MRI studies. Age, sex, and group (HC or Pain) were modelled as covariates during harmonisation to ensure *neuroHarmonize* does not remove the variance associated with these variables.

### Statistical analyses

A series of multiple linear regressions were performed in SPM12 to determine the main effects of group (HC vs Pain), severity of depressive symptoms (BDI total score) and their interaction (the product of group by mean-centred BDI total score) on patterns of seed-based connectivity (one model for each seed). Whole-brain statistical significance was set at an uncorrected voxelwise threshold of *p*<0.001, to which family-wise error correction was applied to the cluster statistics (*pFWEc*<0.05). An additional Bonferroni correction was applied to cluster statistics, to account for the number of seed regions studied (*pFWEc*<0.017).

In case of significant interaction (*pFWEC*<0.017), signal at the peak of the identified clusters was extracted and moderation analyses were formally tested using the ‘interactions’ package (v1.1.5) (Long, 2021) in R (v4.1.2) (R Core Team, 2021) and RStudio (v1.4.1717) (RStudio Team, 2021). First, we tested our a priori hypothesis that the severity of depressive symptoms would moderate the impact of chronic pain on functional connectivity. Second, for sake of completeness, group was also tested as moderating the relationship between the severity of depressive symptoms and patterns of functional connectivity. In addition, within each model, the Davidson-McKinnon correction (HC3) was used to account for potential issues related to heteroskedasticity (Hayes and Cai, 2007), using the R package ‘sandwich’ (Zeileis, 2004, Zeileis *et al*., 2020). Within each significant model, statistical significance was set at a threshold of *p*<0.05.

## Results

### Participant characteristics

Age, sex distribution, and levels of state or trait anxiety as measured by the STAI were not significantly different between the groups. However, the Pain group reported higher levels of depression (BDI total score) compared to the HC group (*Cohen’s d*=0.65). In addition, scanning sites were similarly distributed across the groups [χ^*2*^*(1)=*0.085, *p*=0.798].

### Seed-based functional connectivity

#### Left amygdala

There were no significant main effects of group, depression or their interaction on patterns of functional connectivity with the left amygdala seed region.

#### Right amygdala

There was a significant positive association between the severity of depressive symptoms (BDI total score) and connectivity between the right amygdala and the mPFC [peak MNI coordinates (−8,42,-8), k=743 voxels, *pFWEc*<0.001]. However, this was in the context of a significant association between the group-by-depressive symptom interaction and connectivity between the right amygdala seed region and a cluster largely overlapping the mPFC cluster found associated with depressive symptom severity [peak MNI coordinates (2,42,-10), k=631 voxels, *pFWEc*<0.001]. The moderation analysis was statistically significant [model statistics: adjusted *R*^*2*^=0.250, *F(3,54)*=7.319, *p*<0.001]. When BDI score was entered as moderator (see Figure 1A), connectivity between the right amygdala and the mPFC was weaker in the Pain group compared to the HC group at low levels of BDI scores (*b*=-0.236,*se*=0.096, *t*=-2.446, *p*=0.018, 95%CI[-0.429,-0.043]), but was stronger at high levels of BDI scores (*b*=0.423, *se*=0.125, *t*=3.386, *p*=0.001, 95%CI[0.173,0.674]). There were no group differences in patterns of connectivity between the right amygdala and the mPFC at average BDI scores (*b*=0.094, *se*=0.072, *t*=1.298, *p*=0.200, 95%CI[-0.051,0.239]). When group was entered as moderator (see Figure 1B), the moderation analysis indicated that increasing BDI scores were associated with decreased connectivity between the right amygdala and the mPFC in the HC group (*b*=- 0.034, *se*=0.011, *t*=-2.989, *p*=0.004, 95%CI[-0.057,-0.011]) and with increased connectivity in the Pain group (*b*=0.016, *se*=0.006, *t*=2.637, *p*=0.011, 95%CI[0.004,0.028]).

**Figure 1.**
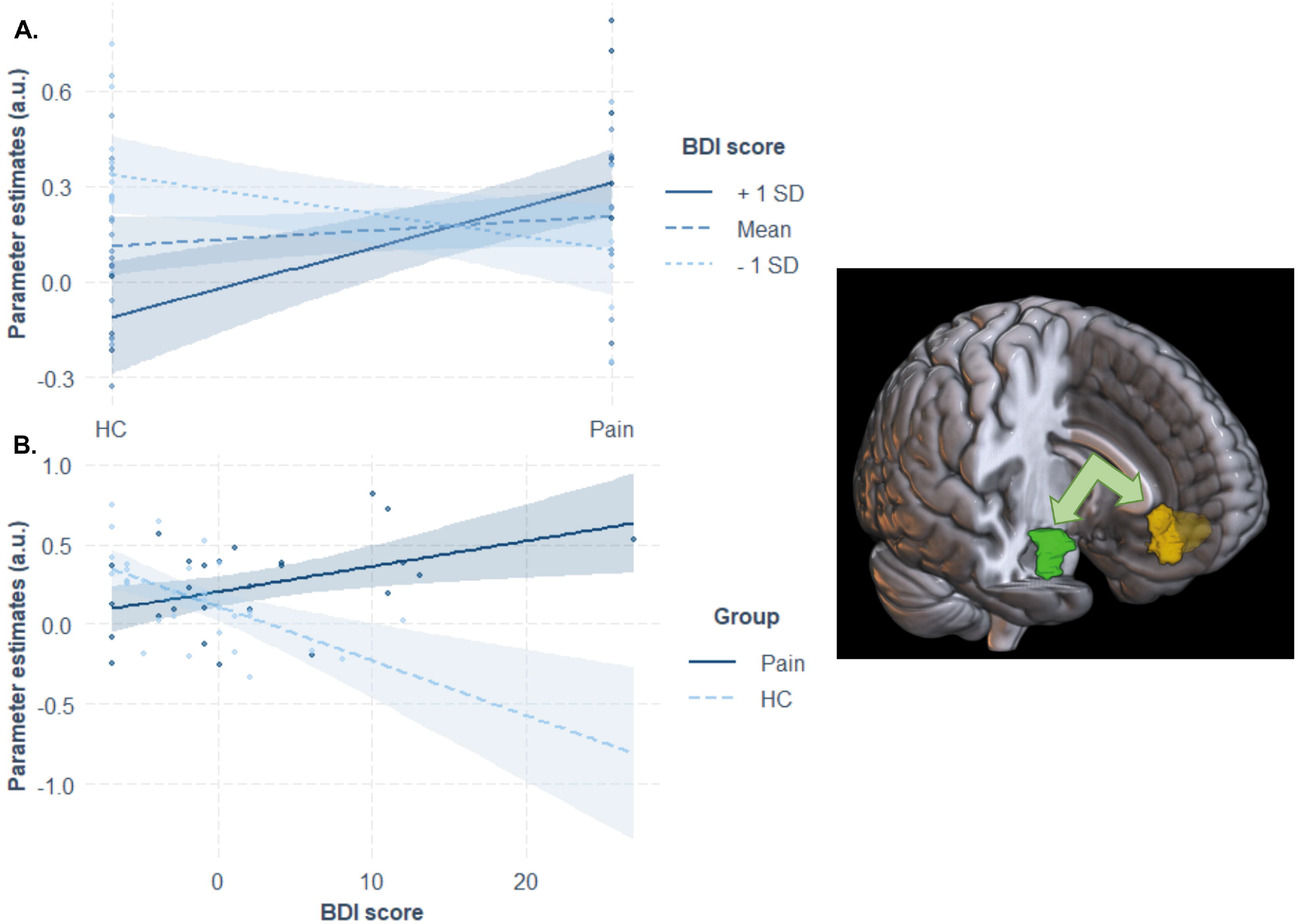
Association between group-by-depressive symptom severity interaction and functional connectivity between the right amygdala seed and the medial prefrontal cortex. Moderation analyses using (A) depressive symptoms, and (B) groups as moderator indicate that: (A) compared to the healthy control group (HC), the Pain group showed weaker connectivity between the right amygdala (in green) and the mPFC (in yellow) at low levels of depressive symptoms (dotted line), but stronger connectivity at higher levels of depressive symptoms (plain line); there was no group difference at average levels of depressive symptoms (dashed line). (B) As levels of depressive symptoms increased, connectivity between the right amygdala and the mPFC decreased in the HC group, and increased in the Pain group

This was also in the context of weak effects of group: compared to the control group, weaker connectivity between the right amygdala seed and the right precentral gyrus [peak MNI coordinates (24,-16,76), k=150 voxels, *pFWEc*=0.042] and stronger connectivity with the pars orbitalis of the right inferior frontal gyrus [peak MNI coordinates (46,32,-8), k=145 voxels, *pFWEc*=0.048] were evident. However, these effects of group failed to survive the adjusted statistical threshold (*pFWEc*<0.017).

#### Medial prefrontal cortex

There was a significant effect of group, with weaker connectivity between the mPFC seed region and the left parahippocampal gyrus [peak MNI coordinates (−24,-20,-30), k=194 voxels, *pFWEc*=0.015; see Figure 2) evident in the Pain group compared to HCs. There were no significant effect of depressive symptoms severity or interactive effects on the connectivity between the mPFC seed and the left parahippocampal gyrus, and there were no significant effect of group, depressive symptoms severity or their interaction with connectivity between the mPFC and any other region in the brain.

**Figure 2.**
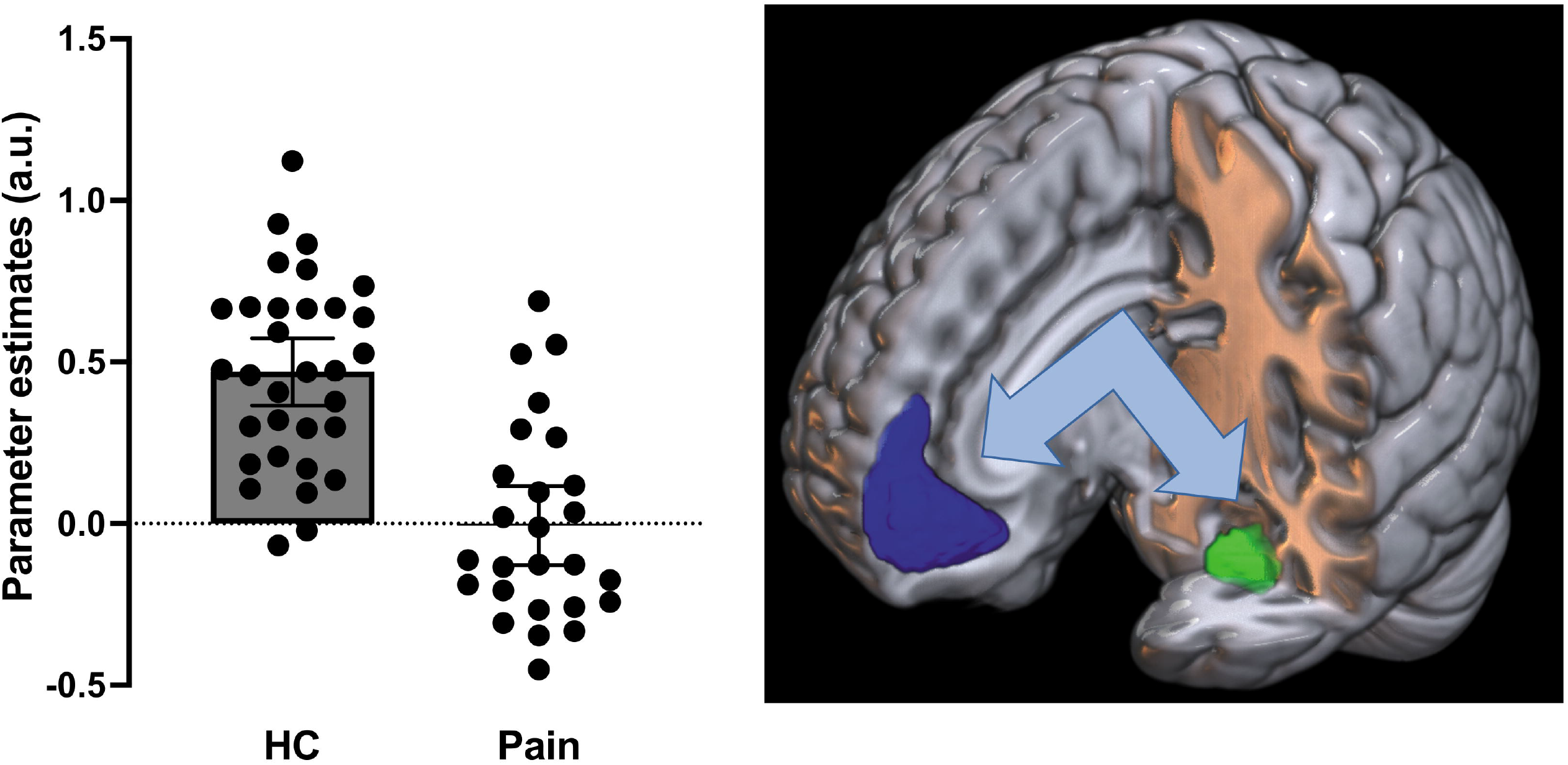
Group difference on patterns of functional connectivity between the mPFC seed and the left parahippocampal gyrus. Compared to the HC group, the Pain group showed weaker connectivity between the mPFC seed (in blue) and the left parahippocampal gyrus (in green)

### Exploratory correlation analyses

Exploratory two-tailed Pearson’s correlations were conducted to rule out potential confounding effects of pain duration and pain intensity (using the pain diary measure) on levels of depression (BDI total score). However, there were no significant associations between pain duration (*r*=-0.035, *p*=0.864) or pain intensity (*r*=0.063, *p*=0.760) and levels of depressive symptoms.

## Discussion

In the context of overall more severe depressive symptoms in the Pain group compared to healthy controls, depressive symptoms moderated the impact of chronic pain on resting-state functional connectivity between regions critical for emotion processing (right amygdala) and regulation (mPFC). In particular, compared to controls, people with chronic pain showed weaker connectivity between the right amygdala seed region and the mPFC at low levels of depressive symptoms, and stronger connectivity at high levels of depressive symptoms. In addition, as levels of depressive symptoms increased, connectivity between these regions decreased in healthy controls and increased in people with pain. Finally, compared to the control group, people with chronic pain showed weaker connectivity between the mPFC and the left parahippocampal gyrus, independently of the levels of depressive symptoms reported.

Partly consistent with our hypothesis, the relationship between the strength of resting-state functional connectivity between the right (but not left) amygdala and the mPFC, and chronic pain (compared to healthy controls), was moderated by the severity of depressive symptoms reported: compared to controls, weaker connectivity was evident at lower levels of depressive symptoms, and stronger connectivity was evident at higher levels of depressive symptoms, in people with chronic pain. As a core node of the DMN, the mPFC specializes in the treatment of affective stimuli (Lieberman *et al*., 2019) and exerts inhibitory top-down control on amygdalar activity (Motzkin *et al*., 2015, Quirk *et al*., 2003). Importantly, the mPFC is also a central hub for both cognitive and affective comorbid states often reported in chronic pain, including depression (Kummer *et al*., 2020). In addition, increased severity of depressive symptoms in the healthy control group was associated with decreasing amygdala-mPFC connectivity strength. This is consistent with previous reports of reduced connectivity between these regions in adults and adolescents with major depressive disorder (Connolly *et al*., 2017, Tang *et al*., 2013, Veer *et al*., 2010), and with reduced top-down control over amygdala activity leading to the mPFC inability to down-regulate negative affect (Johnstone *et al*., 2007). On the other hand, increasing severity of depressive symptoms was associated with increasing strength of connectivity in people with chronic pain. This suggests that the top-down regulation provided by the mPFC to the amygdala may be inefficient, reflecting potential mood-related maladaptive consolidation of aberrant affective information in this population.

Mechanisms by which depressive symptoms impact functional connectivity between the mPFC and amygdala in chronic pain are unclear but may be associated with changes (increase) in peripheral inflammation. Increased levels of peripheral inflammation are common in chronic pain and depression (Beurel *et al*., 2020, Marchand *et al*., 2005), and are considered a common mediator in both conditions (Walker *et al*., 2014). Increased inflammation can be triggered by sustained exposure to psychosocial stressors, such as chronic pain (Abdallah and Geha, 2017, Haroon *et al*., 2012), inducing the release of stress-related glucocorticoids (i.e., cortisol). Released cortisol will in turn trigger glia activation and cytokine production (Jauregui-Huerta *et al*., 2010), that downregulate glutamate levels in the mPFC (Milligan and Watkins, 2009), as observed in both chronic pain (Naylor *et al*., 2019) and depression (Belleau *et al*., 2019), and may result in an inefficient mPFC top-down regulation. Results from the present study indicate that changes in functional connectivity between the mPFC and the amygdala are sensitive to depressive symptoms in individuals with chronic pain. Although these changes are different (opposite direction) in people with chronic pain compared to pain-free individuals, they may operate on a continuum, via common neurobiological pathways including chronic stress, inflammation, and glutamate availability in the mPFC. This mechanistic explanation is plausible, but it remains speculative and future large longitudinal studies integrating markers of inflammation, brain function and neurochemistry, as well as clinical and behavioural phenotypes are necessary to better understand and confirm these mechanisms.

The connectivity between the mPFC seed region and the left parahippocampal gyrus was stronger in the control group compared to the group of individual suffering from chronic pain, independently of the severity of depressive symptoms. This is consistent with a previous study proposing that aberrant connectivity between the hippocampus/parahippocampal gyrus and the mPFC could represent a predictive marker of the transition to chronic pain (Mutso *et al*., 2014). In addition to support hippocampal functions in assimilating and turning new memories in long-term permanent knowledge (Brod *et al*., 2013, Preston and Eichenbaum, 2013, van Kesteren *et al*., 2012), the parahippocampal gyrus is a critical hub at the interface between the mPFC and the hippocampus for processing the spatial (‘where’) context of memories (Preston and Eichenbaum, 2013). Thus, weaker connectivity between the mPFC and the left parahippocampal gyrus could reflect brain function associated with aberrant contextual pain memory, and maybe more broadly, with deficits in long-term memory in chronic pain (Mazza *et al*., 2018). This interpretation is largely speculative, as long-term contextual memory has not been tested in this study.

Contrary to our hypothesis, the connectivity between the mPFC and the PCC/precuneus was not associated with the severity of depressive symptoms. Importantly, our results provide some evidence that the connectivity between the mPFC and PCC/precuneus is not sensitive to the severity of depressive symptoms, but rather associated with a diagnosis of major depressive disorder (Yan *et al*., 2019). We also note that none of our participants was formally diagnosed with major depressive disorder, and that our study may lack of enough statistical power to uncover subtle effects. Future studies including a group of individuals with chronic pain and major depressive disorder is required to disentangle the changes in DMN connectivity, specific and common to chronic pain or depression.

Overall, results indicate that depressive symptoms and emotional brain circuits are potential targets for interventions in chronic pain. For instance, repeated transcranial magnetic stimulation (rTMS) targeting the mPFC could help normalise the top-down control of the mPFC on the amygdala, especially in people reporting higher levels of depression. Significant reduction of symptom severity in depressed individuals, relative to conventional rTMS of the left dorsolateral prefrontal cortex, was reported using mediofrontal double cone coil stimulation of the anterior cingulate cortex (Kreuzer *et al*., 2015). Another study reported reduced depressive symptoms following the stimulation of the right orbitofrontal cortex (at the AF8 site, defined according to the international 10–20 EEG system) (Feffer *et al*., 2018). It is important to note that both stimulation sites are close, but not exactly matching the location of our mPFC cluster. Efficacy of rTMS of the mPFC in reducing depressive symptoms and normalising emotional processing/regulation in people suffering from chronic pain should be considered for future clinical trials. Normalisation of emotion/affect dysregulation can reduce pain severity (Connelly *et al*., 2007, Linton, 2013, Norman-Nott *et al*., 2021, Trucharte *et al*., 2020), and could be key targets to prevent the development and maintenance of chronic pain (Lumley *et al*., 2011). It is also important to note that people with different types of chronic pain (i.e., neuropathic and nociceptive) exhibit comparable negative affective-motivational and cognitive-evaluative states including similar levels of depression (Gustin *et al*., 2011). Thus, targeting emotional/affective processing areas such as the mPFC may be key in reducing both the affective and physical suffering regardless of chronic pain type.

This study has several limitations. First, the studied sample size was relatively small, preventing the identification of smaller effects. Second, people with chronic pain included in this study suffered from different conditions and pain was reported at various locations over the body that may potentially have influenced patterns of brain function. Despite these limitations, the observed effects are strong and are present in a heterogeneous group of pain sufferers, indicating that they may represent a common feature across pain disorders. Replication studies in larger clinical groups are required. Third, most clinical cases were using a variety of medications, mostly analgesics. Although the amount of drugs taken by each individual was not controlled for and may have influenced brain function, it represents an ecological sample of what people suffering from chronic pain generally use.

In conclusion, severity of depressive symptoms moderates resting-state functional connectivity between region critical for emotional recognition (amygdala) and regulation (mPFC) in people with chronic pain and healthy controls. These results may have implications for the choice of treatment for chronic pain, in the context of reported depressive symptoms. In particular, future studies should consider testing the efficacy of rTMS of the mPFC in people suffering from chronic pain. Targeting the mPFC may ameliorate both affective and physical suffering of people with chronic pain.

## Data Availability

All data produced in the present study are available upon reasonable request to the authors

## Acknowledgments

The authors acknowledge the volunteers who participated in this study, and assistance of previous students with data collection and entry and medical personnel with participant recruitment.

